# Expanded Access use of Intravenous Gene Transfer with AAV9-GLB1 in a Juvenile GM1 Gangliosidosis Participant

**DOI:** 10.64898/2026.09.04.26361573

**Authors:** Connor J. Lewis, Hera Akmal, Precilla D’Souza, Jean M. Johnston, Sumaiya Ashraf, Gilbert Vezina, Selby I. Chipman, Meghan Blackwood, Muhammad H. Yousef, Zenaide Quezado, William A. Gahl, Barry J. Byrne, Terence R. Flotte, Xuntian Jiang, Audrey Thurm, Amanda L. Gross, Allison M. Keeler, Douglas R. Martin, Miguel Sena-Esteves, Heather L. Gray-Edwards, Raymond Y. Wang, Maria T. Acosta, Cynthia J. Tifft

## Abstract

GM1 gangliosidosis is an inherited and progressively neurodegenerative lysosomal storage disorder with no approved therapy. We report 5-year safety and 3-year clinical, biochemical, and neuroimaging efficacy outcomes following expanded-access intravenous AAV9-GLB1 gene therapy in GT01, a 6-10-year-old female with juvenile-onset GM1 gangliosidosis. During participation in a natural history study before gene transfer, GT01 developed seizures, dysarthria, loss of ambulation, and progressive neurodegeneration. She received a single intravenous administration of AAV9-GLB1 at 1.5×10^13^ vector genomes per kilogram of body weight. Early improvements included the resolution of dysphagia, increased interactions with others, assisted ambulation, and gains in specific domains of adaptive functioning. She was seizure-free without anti-epileptic drugs 18 months post dosing with sustained seizure freedom at the study completion. Cerebrospinal fluid concentrations of GM1 ganglioside and H3N2b (a pentasaccharide biomarker indicative of biochemical improvement in this disorder) concentrations decreased by 50% and β-galactosidase reached normal activity. Brain, thalamic, and net fiber tract volume assessed by differential tractography increased and ventriculomegaly improved three years post-dosing. These results indicate that meaningful benefit can be achieved even in advanced neurologic disease.

## Introduction

GM1 gangliosidosis (GM1) is an ultra-rare, progressive, and fatal lysosomal storage disorder caused by biallelic pathogenic variants in *GLB1*, encoding β-galactosidase^1^. Deficient β-galactosidase activity leads to the accumulation of GM1 ganglioside primarily within the central nervous system (CNS), where the ganglioside concentration is highest^2^. GM1 is clinically categorized into infantile, late-infantile, juvenile, and late-onset subtypes based on age of symptom onset, symptom severity, and rate of progression, and is generally inversely correlated with enzyme activity^3,4^. Infantile GM1 is the most severe phenotype, characterized by symptom onset before 6 months of age and clinical findings including a cherry red macula, hepatosplenomegaly, developmental delay, and cardiomyopathy with a median survival of 19 months^3^. The late-infantile subtype presents at approximately one year of age and the juvenile subtype presents between the ages of 3 and 5 years^5^. The juvenile disease subtype is characterized by dystonia, dysarthria, seizures, skeletal abnormalities, and developmental delays^4,5^. Patients with late-infantile GM1 typically survive into the first or second decade of life, and patients with the juvenile subtype can live into their fourth decade^4^.

There are currently no approved disease-modifying therapies for GM1. However, recent early-phase clinical data suggest that AAV9-mediated gene therapy is well tolerated and may offer biochemical and neuroimaging improvements or stabilization in individuals with late-infantile and juvenile GM1^6^. These findings stem from the NIH-led AAV9-GLB1 gene therapy trial (NCT03952637), which evaluated intravenous delivery of an AAV9 vector carrying a functional *GLB1* transgene^6^. Other ongoing clinical trials in GM1 include oral substrate reduction therapy (NCT07082543), and two additional gene therapies with one injected into the cisterna magna (NCT04713475) and one involving prenatal treatment (NCT07479953).

Here we present the findings from an expanded access investigation of a participant with advanced juvenile-onset GM1 who was screen failed for the phase 1/2 AAV9-GLB1 gene therapy trial due to low adaptive functioning scores, but who received intravenous AAV9-GLB1 at age 6-10 through an FDA-authorized expanded access pathway. This report provides the first 5-year safety and 3-year clinical, biochemical, and neuroimaging follow-up of AAV9-GLB1 gene therapy in a participant with advanced juvenile GM1 gangliosidosis, offering insight into the safety and potential efficacy of this treatment in the setting of significant pre-existing neurodegeneration.

## Results

### Natural History Study Evaluations

GT01 is a now a 16-20-year-old female with a past medical history of normal development until 0-5 years, when her parents first noted speech delay and unsteady gait. At 0-5 years, she developed stuttering, strabismus, and ataxic gait with falls, marking symptom onset. Exome sequencing identified compound heterozygous variants in *GLB1*, i.e., c.245C>T (p.Thr82Met) and c.367G>A (p.Gly123Arg), biochemical testing showed markedly reduced β-galactosidase enzyme activity (0.1 nmol/min/mg [reference range: 0.5-3.6 nmol/min/mg]), confirming the diagnosis of GM1 gangliosidosis.

During the four years prior to receiving AAV9-GLB1, she was enrolled in a natural history study (NCT00029965) with serial clinical evaluations, electroencephalogram (EEG), magnetic resonance imaging (MRI) including diffusion tensor imaging (DTI), and Vineland Adaptive Behavior Scale evaluations (second edition). During this natural history study period, GT01’s condition worsened steadily. She developed seizures and incontinence, became non-ambulatory, and exhibited behavior difficulties and severe dysarthria, with speech limited to single words. Three years prior to gene transfer, her EEG was normal and MRI showed mild cerebellar atrophy. Two years prior to gene transfer there were rare epileptiform spikes emerged on EEG, coinciding with seizure onset, and anti-epileptic drugs (AEDs) were initiated. MRI demonstrated mild cerebral atrophy and Vineland standard scores also declined. At her baseline evaluation, one month prior to gene transfer, while receiving AEDs, EEG revealed abnormal spiking and mild background slowing without frank epileptiform discharges. MRI demonstrated progressive cerebral and cerebellar atrophy. Her Vineland (third edition)^7^ Adaptive Behavior Composite (ABC) composite standard score was a 33 (maximum score of 140; a score of 40 or more was required), rendering her ineligible for the gene therapy clinical trial (NCT03952637) and prompting expanded-access considerations^6,7^. Her baseline evaluation also yielded a Clinical Global Impression-Severity (CGI-S) score of “5 – Markedly Ill”. Her baseline antibody titer was <1:25 and is a comparative estimate based on data from participants in the phase 1/2 study^6^. She received an intravenous administration of AAV9-GLB1 at a dose of 1.5×10^13^ vector genomes per kilogram of body weight.

### Safety and Immune response

GT01 experienced 13 adverse events (AEs) over the five-year study period including two serious adverse events (SAEs, Table 1, Supplement Table S1). The two SAEs occurred prior to the administration of AAV9-GLB1 and included dehydration and exacerbation of GT01’s underlying seizure disorder requiring readmission 25 days prior to gene transfer. GT01 was given intravenous fluids, and her levetiracetam dose was increased which resolved this event in two days. The other SAE was an injection site reaction at the peripherally inserted central catheter (PICC) site 6 days prior to gene transfer with negative blood cultures that resolved in two days with a dressing change. Both SAEs were deemed not related to AAV9-GLB1 due to their timing. There were three other AEs that occurred prior to the administration of AAV9-GLB1 including vomiting 29 days prior to gene transfer, an infusion-related reaction to rituximab 21 days prior to gene transfer, and mild epistaxis which resolved 13 days prior to gene transfer^6^. There were three AEs deemed possibly related to AAV9-GLB1. On the day of gene transfer she experienced hypotension which did not require intervention and resolved on the same day. On day one following gene transfer, she experienced vomiting and decreased bowel sounds and GT01 was given intravenous fluids, and a bowel regimen was administered. On day seven following gene transfer, GT01 experienced thrombocytopenia with a nadir of 88 K/mcL (reference range: 202-403 K/mcL) and a peripheral smear showing decreased platelets without clumping. There was no evidence of epistaxis, bruising, or additional signs of abnormal bleeding. On day 9 following gene transfer, GT01 was cleared by hematology and her platelet count returned to normal (> 202 K/mcL) on day twelve following gene transfer without intervention. There were five additional AEs that occurred after gene transfer and were deemed unrelated to AAV9-GLB1 including oral mucositis (day 36), gastritis (day 42), a stye (day 90), dental caries (day 107), and a viral illness five months after gene transfer.

**Table 1.**
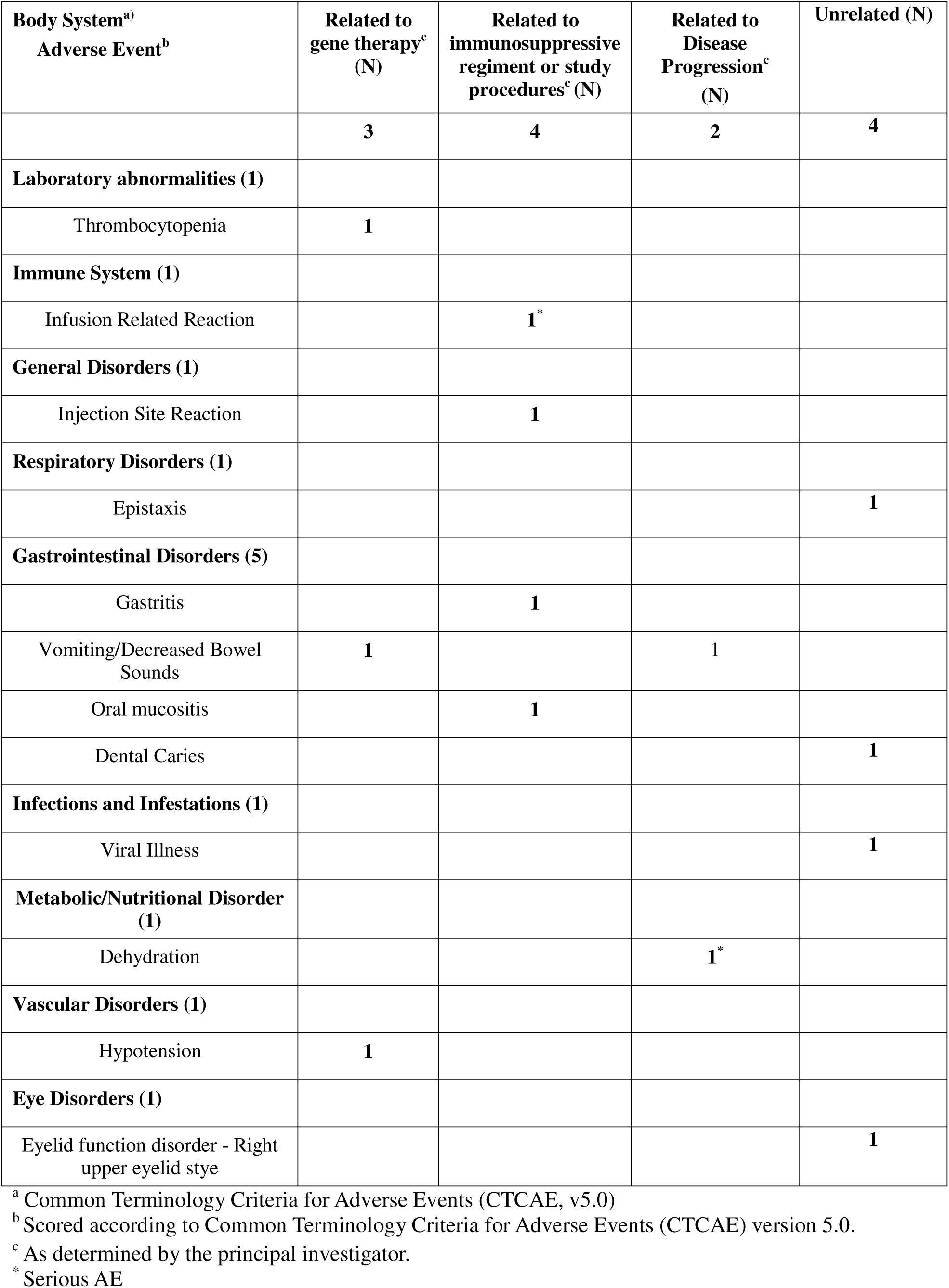
Adverse Events (AE)

| **Body System<sup>a)</sup>**<br>**Adverse Event<sup>b)</sup>** | **Related to**<br>**gene therapy<sup>c)</sup>**<br>**(N)** | **Related to**<br>**immunosuppressive**<br>**regiment or study**<br>**procedures<sup>c)</sup> (N)** | **Related to**<br>**Disease**<br>**Progression<sup>c)</sup>**<br>**(N)** | **Unrelated (N)** |
| --- | --- | --- | --- | --- |
|  | **3** | **4** | **2** | **4** |
| **Laboratory abnormalities (1)** |  |  |  |  |
| Thrombocytopenia | **1** |  |  |  |
| **Immune System (1)** |  |  |  |  |
| Infusion Related Reaction |  | **1\*** |  |  |
| **General Disorders (1)** |  |  |  |  |
| Injection Site Reaction |  | **1** |  |  |
| **Respiratory Disorders (1)** |  |  |  |  |
| Epistaxis |  |  |  | **1** |
| **Gastrointestinal Disorders (5)** |  |  |  |  |
| Gastritis |  | **1** |  |  |
| Vomiting/Decreased Bowel<br>Sounds | **1** |  | **1** |  |
| Oral mucositis |  | **1** |  |  |
| Dental Caries |  |  |  | **1** |
| **Infections and Infestations (1)** |  |  |  |  |
| Viral Illness |  |  |  | **1** |
| **Metabolic/Nutritional Disorder**<br>**(1)** |  |  |  |  |
| Dehydration |  |  | **1\*** |  |
| **Vascular Disorders (1)** |  |  |  |  |
| Hypotension | **1** |  |  |  |
| **Eye Disorders (1)** |  |  |  |  |
| Eyelid function disorder - Right<br>upper eyelid stye |  |  |  | **1** |

<sup>a</sup> Common Terminology Criteria for Adverse Events (CTCAE, v5.0)<sup>b</sup> Scored according to Common Terminology Criteria for Adverse Events (CTCAE) version 5.0.
<sup>c</sup> As determined by the principal investigator. \* Serious AE

Consistent with the natural history of juvenile GM1 gangliosidosis^5^, GT01’s AST was at the upper limit of normal at baseline (41 U/L) and was further elevated up to 2-fold following gene transfer; it returned to normal 1 year following gene transfer (Figure 1A). ALT was at the upper limit of normal at baseline (30 U/L), was briefly elevated one week prior to gene transfer (34 U/L) and 10 to 12 weeks post gene transfer but was otherwise normal throughout the study (Figure 1B). GGT was not elevated throughout the study (Figure 1C). As previously described, platelet levels were depleted following gene transfer reached a low level of 88 K/mcL 1 week following gene transfer (Figure 1D). D-dimer was not elevated following gene transfer (Figure 1E), but this test was performed less frequently compared to our previous study^6^. Total bilirubin was not elevated during the study (Figure 1F), and alkaline phosphatase was transiently elevated at one year post gene transfer (Figure 1G).

**Figure 1.**
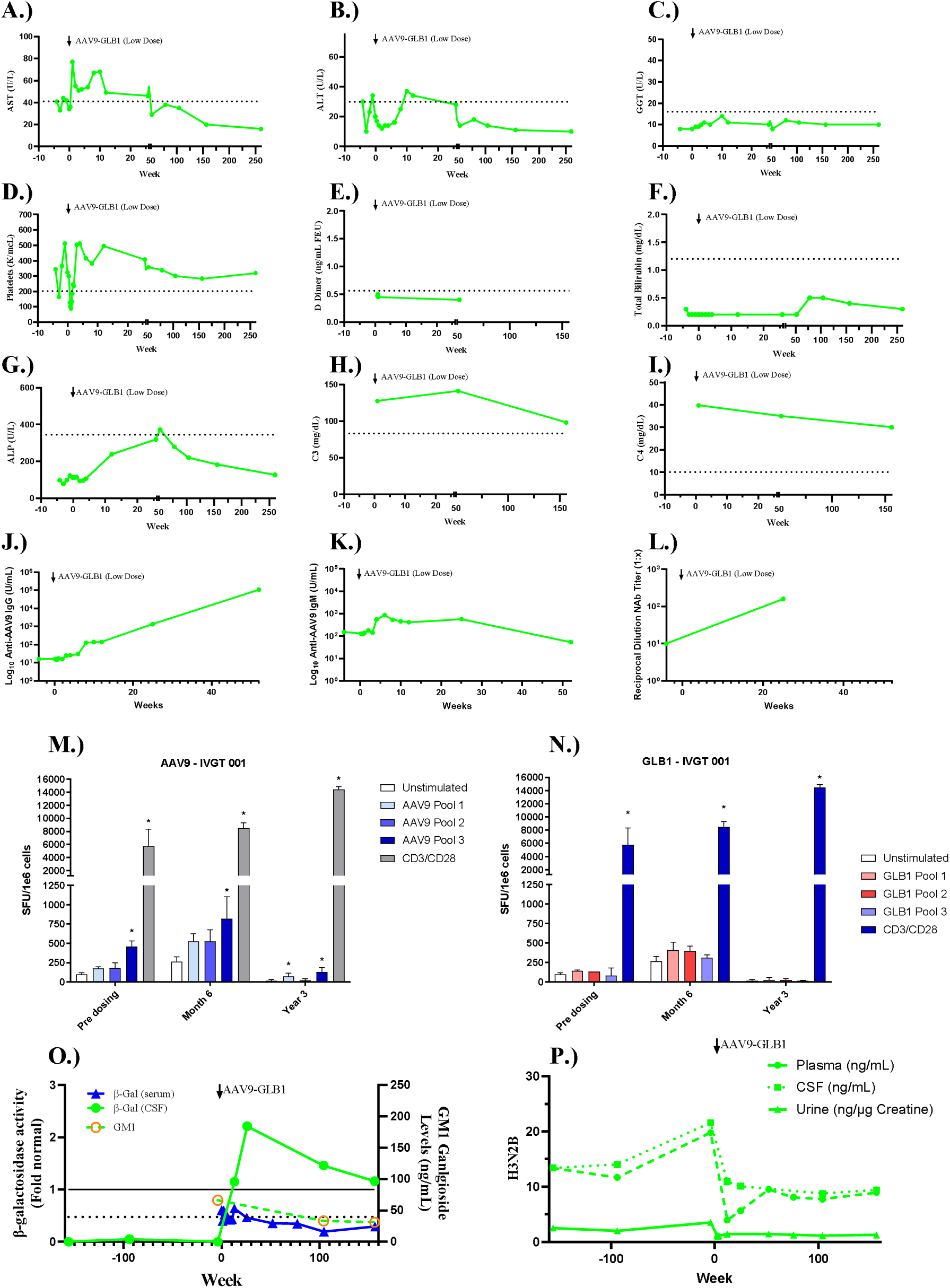
Biochemical Outcomes Following AAV9-GLB1. A.) Aspartate aminotransferase (AST) in serum; the dotted line represents the upper limit of normal (ULN, 41 U/L). B.) Alanine aminotransferase (ALT) in serum (ULN = 30 U/L). C.) Gamma-Glutamyl Transferase (GGT) in serum (ULN = 16 U/L). D.) Platelet count in serum; the dotted line represents the lower limit of normal (LLN, 202 K/mcL). E.) D-Dimer level in serum (ULN = 0.56 ng/mL FEU). F.) Total bilirubin levels in serum (ULN = 1.2 mg/dL). G.) Alkaline phosphatase in serum (ULN = 345 U/L). H.) Complement C3 levels in serum (LLN = 83 mg/dL). I.) Complement C4 levels in serum (LLN = 10 mg/dL). J.) Anti-AAV9 IgG levels in serum. K.) Anti-AAV9 IgM levels in serum. L.) Neutralizing antibodies to the AAV9 capsid in serum. M.) Capsid specific ELISpots, IFN-γ ELISpot results for capsid specific immune responses. Positive responses designated by * and determined if they were ≥ 50 SFU/1e6 cells and at least 3X unstimulated negative control. CD3/CD28 stimulation was used as positive control. Positive and negative controls were run on all participants at all timepoints. N.) Transgene specific ELISpots, IFN-γ ELISpot results for capsid specific immune responses. Positive responses designated by * and determined if they were ≥ 50 SFU/1e6 cells and at least 3X unstimulated negative control. CD3/CD28 stimulation was used as positive control. Positive and negative controls were run on all participants at all timepoints. O.) β-galactosidase activity in (green, closed circles) and GM1 concentration (green, open circles) in CSF and β-galactosidase activity in serum (blue, triangles). The dotted line represents the upper limit of normal for GM1 ganglioside concentration in CSF (39.9 ng/mL at 1 standard deviation above normal^8^). The black filled in line represents normal β-galactosidase activity for both serum and CSF. P.) H3N2b levels in plasma (blue circles), CSF (green squares), and urine (red triangles) calculated relative to the cut-off values from Kell et al^53^. The dotted line in all figure panels indicate upper or lower limits of normal.

Complement C3 and C4 were not depleted following gene transfer (Figure 1H and 1I), but these tests were performed less frequently compared to our previous study^6^. Anti-AAV9 IgG peaked one year following gene transfer (most recent collection, Figure 1J). Anti-AAV9 IgM peaked 6 weeks following gene transfer and returned to baseline 1 year following gene transfer (Figure 1K). Anti-AAV9 neutralizing antibodies were elevated following gene transfer, but with only two timepoints collected, it is not possible to determine the peak (Figure 1L). GT01 had a positive IFN-γ-ELISpot to the AAV capsid at baseline that remained positive at 6 months and 3 years following gene transfer (Figure 1M). Transgene-specific immune responses were not observed (Figure 1N).

### Biochemical Response to Gene Transfer

CSF β-galactosidase activity in CSF was assessed two and three years prior to enrollment in the expanded access clinical trial. CSF β-galactosidase activity was 0 percent of normal three years prior and 5 percent of normal two years before gene transfer (Figure 1O). GT01’s CSF β-galactosidase activity peaked at 2.21 times normal activity 6 months following gene transfer and remained above normal 3 years following gene transfer (Figure 1O). CSF GM1 ganglioside levels were 66.1 ng/mL at baseline (upper limit of normal = 39.9 ng/mL), 33.2 ng/mL 2 years following gene transfer, and 31.1 ng/mL 3 years following gene transfer (Figure 1O)^8^. H3N2b levels were analyzed two and three years prior to gene transfer in urine, serum, and CSF, as an indicator of biochemical response reflective of β-galactosidase activity^9^. H3N2b levels were increased in all three bodily fluids in the 3 years prior to receiving AAV9-GLB1 relative to the normal cutoffs (Figure 1P). CSF, plasma, and urine H3N2b levels decreased in all three biological fluids following gene transfer and remained below baseline 3 years following therapy (Figure 1P). Serum β-galactosidase activity was not elevated in any of the thirteen samples collected post gene transfer compared to baseline despite the elevations observed in CSF and elevations observed in the phase 1/2 trial (Figure 1O)^6^.

### Clinical Evaluations and Outcomes Following Gene Transfer

At 30 days, CGI-I was “3 – Minimally Improved”. She ambulated with assistance, showed increased muscle tone in extremities, and remained alert and interactive. Three months after gene transfer, there was significant improvement in GT01’s ability to eat safely and remained at the study endpoint. At 6 months, Vineland growth scale values (GSV), a longitudinal within-person change measurement^10^, were increased across all domains (Figure 2). She walked slowly with a walker (an improvement compared to a lack of ambulation at baseline), maintained head and trunk control, and exhibited sustained attention and effortful communication attempts.

**Figure 2.**
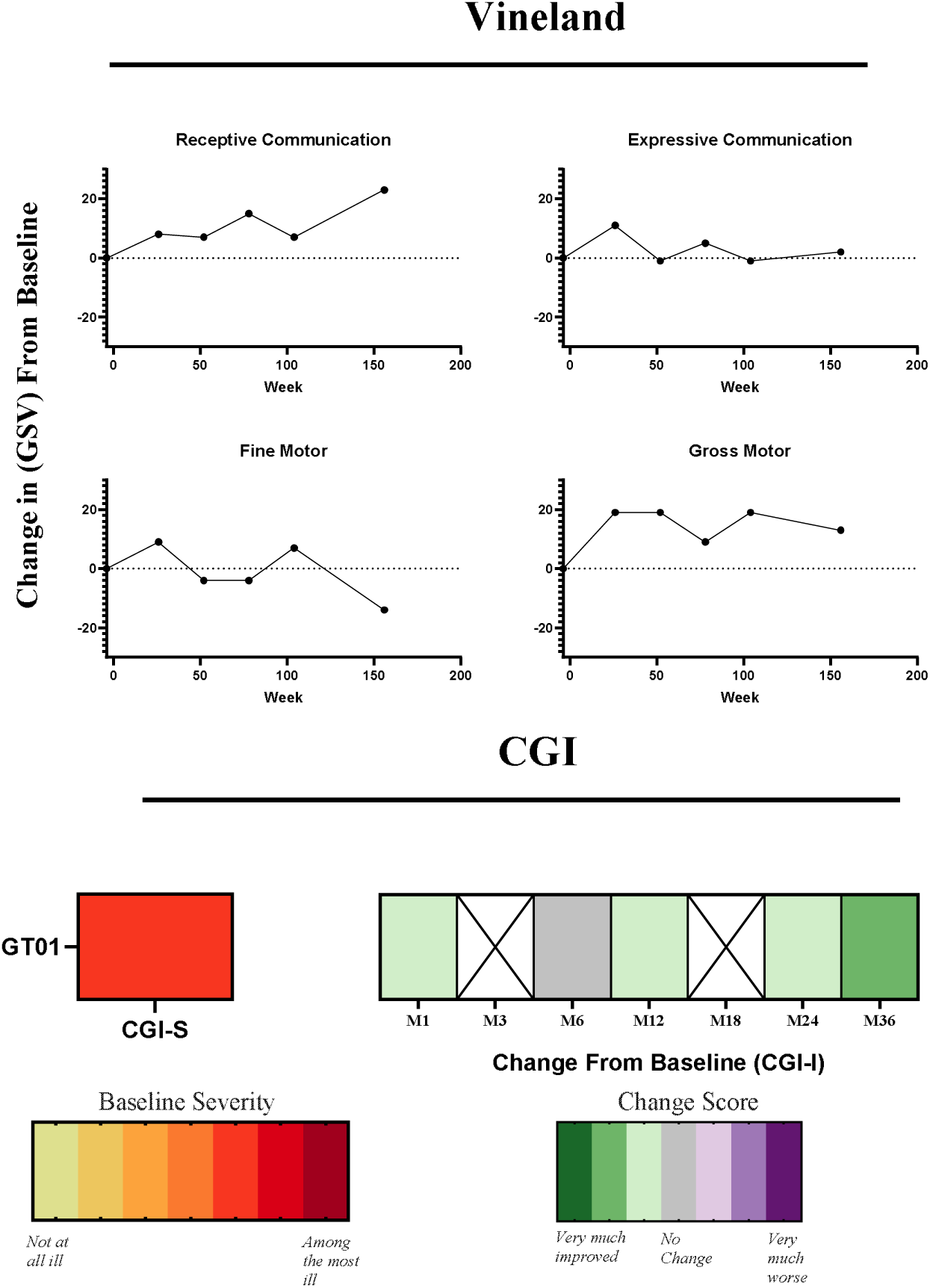
Clinical Outcome Assessments. Vineland-3 Growth Scale Value (GSV) scores presented as a change from baseline for receptive communication, expressive communication, gross motor, and fine motor function. Vineland GSV are shown in supplement figure S1. Clinical Global Impression– Improvement (CGI-I) scores showed meaningful improvement during the first three years after gene therapy, reflecting enhanced engagement, ambulation with assistance, and improved attention. CGI-I was not evaluated at 3 months or 18 months after gene transfer as the family declined in-person follow-up.

One year following gene transfer, Vineland receptive communication GSVs were increased by the same amount as at 6 months, expressive communication values had decreased to baseline levels, gross motor scores remained increased by the same amount as at 6 months, and fine motor skills had decreased to four points below baseline (Figure 2). By 18 months, AEDs were discontinued with no epileptiform abnormalities on EEG, which still demonstrated mild to moderate background slowing. Vineland receptive communication GSV were 15 points above baseline, expressive communication was 5 points above baseline, gross motor scores were 9 points above baseline, and fine motor scores were the same as the previous measurement.

At the 2-year follow-up evaluation, GT01 showed minimal but stable improvement from baseline. Gait ataxia was reduced (an improvement) and she was still able to ambulate with a walker; although she preferred crawling for mobility. This could be attributable at least in part due to her 19.5 cm growth and 8.3 kg weight gain compared to baseline, which significantly changed her center of gravity^11^. Hand use was limited with predominantly closed fists. CGI-I was scored “3 – Minimally Improved” (Figure 2). At two years post gene transfer, cohort average Vineland GSV change from baseline in the phase 1/2 trial were: receptive communication = -5.2; expressive language = -4.6; fine motor = -12.6; and gross motor = -7.2^6^. In contrast, two years post gene transfer, GT01 had GSVs of receptive communication = 7, expressive communication = -1, fine motor = 7, and gross motor = 19.

At her 3-year follow-up evaluation, her parents reported improved attention, social interactions, energy levels, hand-eye coordination, eye gaze and reduced strabismus. CGI-I was scored “2 – Much Improved” (Figure 2). At three years post gene transfer, cohort average Vineland GSV changes from baseline in the phase 1/2 trial were: receptive communication = -1.7; expressive language = -4.7; fine motor = -15.9; and gross motor = -7.6^6^. In contrast, at three years post gene transfer, GT01 had GSVs of receptive communication = 23, expressive communication = 2, fine motor = -14, and gross motor = 13.

### Neuroimaging

Prior to gene transfer, GT01 had T1-weighted MRI scans conducted at ages 0-5 and 0-5 years (prior to enrollment in the natural history study), ages 6-10 and 6-10 (during the natural history study), and at age 6-10 (baseline for expanded access trial). GT01’s rate of whole brain and thalamic atrophy and associated enlargement of the lateral ventricles was consistent with that of the natural history participants (Figures 3A, 3C, and 3E). Three years following gene transfer, whole brain volume increased, thalamic volume increased, and lateral ventricle volume decreased (Figures 3A, 3C, and 1E). This result was unaffected when controlling for intracranial volume (ICV, Supplement Figure S4).

**Figure 3.**
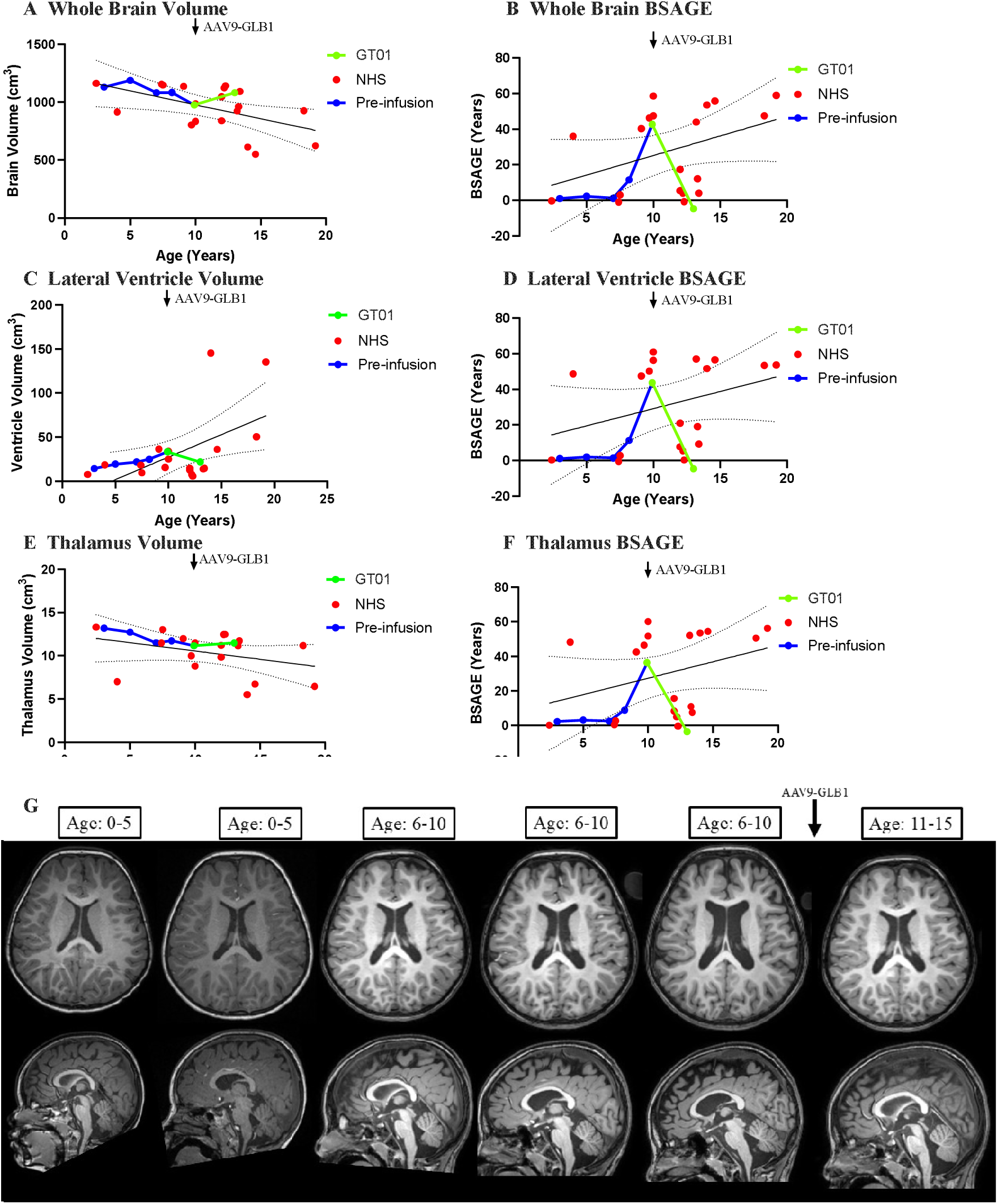
Brain MRI and Volumetric analysis. A.) GT01’s total brain volume from 0-5 - 11-15 pre-infusion (blue) and post-infusion (green) in relation to the baseline scans from untreated natural history participants (red). B.) GT01’s predicted brain structures age gap estimation (BSAGE) for the whole brain from age 0-5 - 11-15 pre-infusion (blue) and post-infusion (green) in relation to the baseline scans from untreated natural history participants (red). C.) GT01’s lateral ventricle volume from age 0-5 - 11-15 pre-infusion (blue) and post-infusion (green) in relation to the baseline scans from untreated natural history participants (red). D.) GT01’s predicted BSAGE for the lateral ventricles from age 0-5 - 11-15 pre-infusion (blue) and post-infusion (green) in relation to the baseline scans from untreated natural history participants (red). E.) GT01’s thalamic volume from age 0-5 - 11-15 pre-infusion (blue) and post-infusion (green) in relation to the baseline scans from untreated natural history participants (red). F.) GT01’s predicted BSAGE for the thalamus from age 0-5 - 11-15 pre-infusion (blue) and post-infusion (green) in relation to the baseline scans from *n* = 18 untreated natural history juvenile GM1 participants (red)^6^. For A-F, a simple linear regression was performed for the cross-sectional untreated natural history study participants’ MRI data with a 95 percent confidence interval shown. G.) Serial MRI shown over the course of the natural history study, showing progressive ventricle enlargement from age 0-5 – 6-10 and a reduction in size at age 11-15 following gene transfer (arrow). Volumes corrected for total ICV and predicted brain ages uncorrected for biological age are included in supplemental figure S4.

In a natural history study of untreated GM1 patients^12^, the rate of brain aging, an imaging marker of neurodegeneration in juvenile GM1 patients was assessed to be more than twice that of controls. Furthermore, the brain structures age gap estimation (BSAGE), i.e., the relative deviation between the predicted age and biological age, was also significantly elevated in juvenile GM1 patients^12^. From age 0-5 – 6-10 years, GT01’s predicted brain age and BSAGE for the whole brain, lateral ventricles, and thalamus were not significantly elevated compared to normal (Figure 3B, 3D, and 3E, Supplement Figure S4). Between the ages of 6-10 and 6-10 years, predicted brain age and BSAGE drastically increased, coinciding with clinical impairment. Similarly to the volumetric analysis, predicted brain age and BSAGE for the thalamus, lateral ventricles, and whole brain decreased drastically at the three-year follow-up post gene transfer (Figure 3B, 3D, and 3E and Supplement Figure S4).

Differential tractography in the natural history of type II GM1 patients (including GT01) has demonstrated consistent declines in fiber tract count and fiber tract volume (Figure 4A, 4B, and 4D)^13^. As shown in Figure 4A, GT01 had a net fiber tract count decline of 1.2 percent and a net fiber tract volume decline of 2.2 percent between the ages of 6-10 and 6-10 years in the natural history study. After receiving AAV9-GLB1, GT01 had significant gains in fiber tracts as shown in Figure 4A. GT01 had a net fiber tract count increase of 3.4 percent and net fiber tract volume increase of 4.8 percent compared to an average of a 0.5 ± 0.2 percent count increase and 1.3 ± 0.5 percent volume increase in the phase 1/2 trial participants evaluated at three years post gene transfer (Figure 4C and 4E)^6^.

**Figure 4.**
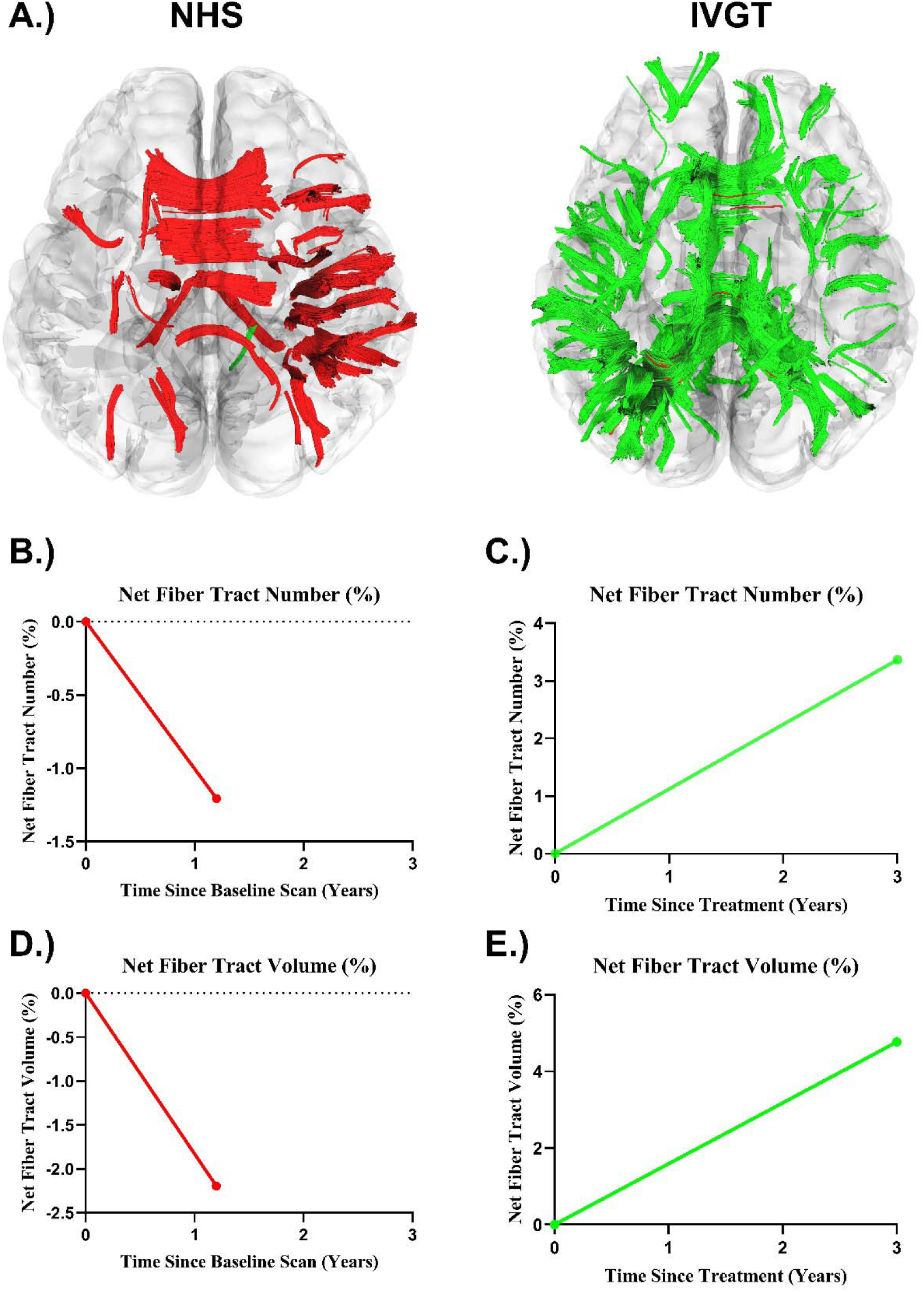
Differential Tractography. **A.)** Differential fiber tractography assessed fiber tract gains (green) and fiber tract losses (red). On the left is GT01’s change in fiber tracts between the ages of 6-10 and 6-10 years old under the natural history protocol. On the right is GT01’s change in fiber tracts between the ages of 6-10 and 11-15 after receiving AAV9-GLB1. B.) Net fiber tract count during the natural history study period. C.) Net fiber tract count during the gene therapy study period. D.) Net fiber tract volume during the natural history study period. E.) Net fiber tract volume during the gene therapy study period.

## Discussion

This study provides further support that intravenous AAV9-GLB1 gene therapy is generally well tolerated, with only mild AEs and can provide sustained clinical benefit in children with Type II GM1 gangliosidosis even when administered at an advanced disease stage^6^. It supports the biochemical and neuroimaging results described in the phase 1/2 trial and preclinical studies^6,14–16^. In addition, this study indicates that the administration of AAV9-GLB1 can result in improved mobility and an arrest of seizures in GM1 patients, a finding observed in preclinical studies but not the phase 1/2 trial^6,16–18^.

There were 13 AEs observed during this investigation; but only three were related to AAV9-GLB1 and they were relatively mild. There were two SAEs that occurred prior to gene transfer, as defined due to requiring hospitalizations, i.e., dehydration resulting in worsening of the patient’s seizure disorder and the other being redness at the PICC site which resolved in two days. The thrombocytopenia observed in this study has been reported in other AAV trials^19^; while thrombocytopenia can be an early sign of thrombotic microangiopathy (TMA), the lowest platelet count observed in this patient (88 K/mcL) is considered Grade 1 (Mild) under Common Terminology Criteria for Adverse Events Grade 1, and there were no physical signs of abnormal bleeding. Furthermore, in gene therapy trials of spinal muscular atrophy, similar transient declines in platelet levels were observed following the administration of AAV9 and quickly return to baseline levels^20^. Ultimately, platelet counts should be closely monitored following the administration of AAV based therapeutics. The overall safety profile of AAV9-GLB1 in this study is similar to that observed in our previous study^6^; vomiting and elevation of liver enzymes were observed following gene transfer but there was no evidence of acute respiratory destress syndrome (ARDS), cholestatic liver failure progressing to sepsis and multiorgan dysfunction, hemophagocytic lymphohistiocytosis (HLH), or cardiogenic shock which have led to fatalities in AAV trials^21–25^.

Biochemical evaluations showed preliminary evidence of efficacy. There were 50 percent reductions in both GM1 ganglioside and H3N2b in CSF and GM1 in CSF reached normal levels^8^. β-galactosidase in CSF reached normal activity 90 days after treatment and remained above normal at 3 years post gene transfer. H3N2b storage was also drastically reduced in plasma and urine. The increased β-galactosidase activity in CSF, decreased GM1 storage in CSF, and decreased H3N2b in CSF, plasma, and urine were all observed in the phase 1/2 trial^6^ but the results are more compelling in GT01; the basis for this is unclear. In contrast, serum β-galactosidase activity remained low in GT01 throughout the study, a finding not observed in the phase 1/2 trial^6^. However, GT01 was also the only patient with a pre-existing T cell immunity to the AAV capsid prior to dosing. Interestingly, several patients within the phase 1/2 trial had decreased serum β-galactosidase activity that corresponded to secondary capsid specific interferon-γ ELISPOT responses and the return of peripheral B-cells after rituximab treatment^6^. This suggests that capsid specific T cell responses could be responsible for loss of peripheral serum β-galactosidase activity, although GM1 ganglioside concentration and β-galactosidase activity in the CSF of GT01 remained stable.

Neuroimaging evaluations demonstrated progressive atrophy of the whole brain and thalamus with associated ventriculomegaly during GT01’s enrollment in the natural history study, particularly between the ages of five and ten. In her first scan post gene transfer (3-year follow-up), total brain and thalamic volume were increased, with shrinkage of the lateral ventricles. The natural history of brain volumetrics in GM1 gangliosidosis has been extensively described^1,5,12,26–28^: the juvenile phenotype, while less severe than the late-infantile phenotype, presents with atrophy of the whole brain, thalamus, cerebellum, corpus callosum, caudate, and lentiform nucleus with associated enlargement of the lateral ventricles. In the phase 1/2 trial^6^, at three years post gene transfer, 2 of the 5 juvenile participants had improvements in the rate of whole brain and thalamic atrophy and 4 of the 5 juvenile participants had improvements in the rate of ventricular enlargement compared with the natural history. In contrast, GT01 had increased whole brain and thalamic volume with reduced lateral ventricular volume suggesting growth of brain parenchyma, not just a reduction in atrophy. A novel imaging marker, BSAGE, has been used in neurodegenerative diseases including multiple sclerosis, Alzheimer’s disease, frontotemporal dementia and other neurodegenerative diseases^29,30^ and was recently described in Type II GM1 gangliosidosis patients^12^ showing that neurodegeneration can resemble brain changes similar to that of older participants^31^. GT01 exhibited accelerated brain aging and increased BSAGE during the same period as atrophy was observed (Figure 3, Supplement Figure S4). After gene transfer, BSAGE and brain age both decreased for the whole brain, thalamus, and lateral ventricles, showing that predicted brain age and BSAGE are reflective of volumetric changes for gene therapy.

In addition, neuroimaging evaluations with differential tractography demonstrated declining white matter fiber tracts during GT01’s enrollment in the natural history study, consistent with previously reported untreated GM1 patients^13^. Volumetric and differential tractography assessed changes in Type II GM1 patients; both have previously been shown to reflect changes in patients’ clinical presentation^13,32^. During the natural history study period, GT01 exhibited declining clinical function, consistent with the volumetric- and differential-tractography assessed changes over the same period. After receiving AAV9-GLB1, GT01 had significant white matter improvement as evident by net fiber tract count and volume gains three years after infusion, in contrast with untreated Type II GM1 patients^13^. When compared to the phase 1/2 trial, GT01 had greater improvement than the rest of the cohort at the three year mark^6^. Neuroimaging changes corresponded to the clinical improvement observed in GT01 over the same period, indicating that AAV9-GLB1 can improve neuroradiologic outcomes in Type II GM1 patients and providing further support that volumetric, differential tractography, and predicted brain age analyses are neuroimaging markers reflective of clinical changes in this population^12,13,32^.

In the three years prior to receiving AAV9-GLB1, GT01 was experiencing a rapid decline in function including the onset of seizures, cerebellar related speech difficulties, and the loss of ambulation. During the first three years after treatment, GT01 experienced meaningful functional gains including the ability to eat safely, assisted with ambulation, improved attention and interactivity, and sustained seizure-freedom after discontinuation of anti-epileptic medications. These functional improvements are compelling, since seizures, mobility, feeding, and interactivity are considered by caregivers of both late-infantile and juvenile patients to be critically important symptoms to treat^33^. Seizures are a prototypical feature of GM1. Although they are more common in the infantile (Type I) and late-infantile (Type IIa) subtypes; when present in the juvenile (Type IIb) subtype, they are persistent and require anti-epileptics^5^. To the best of our knowledge, no study investigating a therapeutic for GM1 has reported seizures stopping after initial onset without anti-epileptic medication. Improvements in EEG were described in the preclinical GM1 feline model following gene therapy^17^. Interestingly, the severity of seizures and the ability to maintain oral feeding were also the most sensitive clinical indicators of stabilization or improvement in a phase 1/2 study of a dual AAVrh8 gene therapy for GM2 gangliosidosis^34,35^. That report emphasized that these two outcomes had been rated as highly important by caregivers in a survey conducted as part of an FDA-sponsored patient-focused drug development meeting^35^. This suggests the importance in evaluating seizures, EEG, and oropharyngeal function as outcomes in Type II GM1 clinical trials.

The clinical benefit of AAV9-GLB1 was also evident by CGI-I scores and Vineland GSVs. CGI-I scores increase (worsening clinical severity) longitudinally in Type II GM1 gangliosidosis patients, reflecting neurodegeneration^32^. In contrast, GT01 had decreasing CGI-I scores, with a CGI-I score of 2 (much improved) three years following gene transfer, indicating that she was clinically improved compared to baseline. Similarly, Vineland scores have been shown to decrease longitudinally in Type II GM1 patients^5^. While gross motor GSVs were consistent with the phase 1/2 trial three years following gene transfer, receptive and expressive language, and fine motor GSVs were between 7 and 25 points higher than the average in the phase 1/2 trial^6^. This indicates clinical and functional improvement in this patient, a significant result, particularly in view of the universal downward clinical and functional trajectory observed in Type II GM1 patients^5,36^.

At her baseline evaluation, GT01 had the most advanced disease compared to participants in the phase 1/2 trial as evident by her baseline CGI-S and Vineland adaptive behavior scores, which made her ineligible for the phase 1/2 trial. In gene therapy studies of neurodegenerative diseases, there is a widespread consensus, based on results of pre-clinical and clinical trials, that the earlier treatment is administered, the better the outcome^37–40^. The results of this study challenge that notion: since GT01 had a stronger response to AAV9-GLB1 clinically and biochemically compared to many of her less severely affected peers^6^. While this result is likely not applicable in all cases, it suggests meaningful benefit is possible despite greater disease severity.

This study had a small sample size (*n* = 1) and open-label design, limiting the generalizability of these results. Furthermore, this study is limited by incomplete data collection; the year 1, 18 months, and year 2 in-person follow-up evaluations were completed via telemedicine due to a worldwide pandemic. This limited the collection of MRI, CSF, and some clinical laboratory assessments at those time points. There were also inconsistencies in T1-weighted MRI (age 0-5 and 0-5 scans) and DTI acquisition protocols; the natural history study had 30 diffusion directions while the expanded access study had 15 diffusion directions.

Taken together, these findings support the results of our previous report of nine Type II GM1 patients showing that AAV9-GLB1 is not associated with significant adverse events^6^. The results of this study demonstrate the ability of AAV9-GLB1 to not only stabilize declining clinical function but also provide potential improvements in the form of seizure cessation and mobility improvements which were not appreciated in our previous report^6^. The study also supports the ability of this therapeutic to modify disease trajectory in the form of biochemical and neuroradiologic improvements in those with more advanced juvenile GM1 gangliosidosis.

## Methods

### Study Design and Enrollment

GT01 was originally enrolled in NCT00029965, the “Natural History of Glycosphingolipid Storage Disorders and Glycoprotein Disorders” at age 6-10, an observational study. At age 6-10, GT01 was evaluated for inclusion into NCT03952637, “A Phase 1/2 Study of Intravenous Gene Transfer With an AAV9 Vector Expressing Human Beta-galactosidase in Type I and Type II GM1 Gangliosidosis” but did not meet inclusion criteria due to Vineland-3 Adaptive Behavior composite standard score of 33 ( 40 needed for inclusion). This expanded access study was created with the intention to treat GT01. The National Institutes of Health Institutional Review board reviewed all three protocols (02-HG-0107, 19-HG-0101, and 19-HG-9958) and the Food and Drug Administration approved both the phase 1/2 trial and expanded access trials (IND 18831). The family gave written informed consent for all three studies in accordance with the Declaration of Helsinki.

### Immunosuppression and AAV9-GLB1 Dose

GT01 received the same immunosuppression regiment as NCT03952637 as described in Lewis et al^6^. Three weeks prior to gene transfer, GT01 received daily rapamycin (0.5-1 mg/m^2^) and continued until 6 months after gene transfer. The dosage of rapamycin was adjusted to maintain a serum trough level of 7-12 ng/mL. Rituximab was also started three weeks prior to gene transfer as a weekly dose of 375 mg/m^2^, and GT01 received 4 doses of rituximab with the last dose given the day before gene transfer. GT01 received intravenous methylprednisolone 1 mg/kg 1-2 h prior to administration of AAV9-GLB1 gene transfer. Oral glucocorticoids in the form of oral prednisolone were given daily starting on day one and ending after day three following gene transfer. GT01 received an intravenous administration of AAV9-GLB1 at a dose of 1.5×10^13^ vector genomes per kilogram of body weight and a rate of 1 mL per minute, consistent with the low-dose cohort^6^. With a dosage weight of 25.6 kg, her absolute vector dosage was 3.84×10^14^ vector genomes.

### Outcomes and Endpoints

This expanded access study did not have any predefined inclusion or exclusion criteria nor specified end points. To this end, where applicable, we evaluated biochemical, clinical, and neuroimaging efficacy data in the same manner as described in our previous report^6^. Safety data including clinical laboratory assessments and adverse events were reported over the entire study duration (five-year period after gene transfer). Adverse events and serious adverse events were assessed by site investigators and reviewed quarterly by a data safety monitoring board. Follow-up assessments were conducted at 30 days post treatment, and annually through 5 years (with years 1, 18-month, year 2, and year 4 performed remotely due to COVID-19 pandemic safety related travel concerns). Evaluations included Clinical Global Impression-Improvement (CGI-I) scoring, Vineland testing, EEG, MRI, DTI, and CSF GM1 ganglioside levels and β-galactosidase activity. DTI data were analyzed using differential tractography to assess longitudinal white matter microstructure changes^41^. A summary of the data collected in this study is included in supplemental table S2.

### Biochemical Assessments

Biochemical assessments were performed as described in Lewis et al.^6^, with clinical laboratory assessments primarily performed at the NIH Clinical Center. ELISpot neutralizing antibody analysis of serum was performed at the University of Massachusetts Chan Medical School (ALK) as described in Flotte et al^34^. GM1 ganglioside concentration in CSF was analyzed by Pharmaron Lab Services LLC (Exton, PA) using a validated liquid chromatography-tandem mass spectrometry (LC-MS/MS) assay as described in Lewis et al^6^. β-galactosidase activity in CSF was analyzed by Auburn University (ALG) as previously described^6^. Serum β-galactosidase activity was analyzed by Greenwood Genetic Center (Greenwood, SC). H3N2b levels were analyzed by Washington University Metabolomics Core (XJ) and analyzed in urine, serum, and CSF using a LC-MS/MS^9^. Baseline Anti-AAV9 antibody titer, IgG, and IgM were analyzed by the University of Florida (BJB) using an ELISA as previously described^6,42^. Viral shedding studies were not conducted for GT01.

### Clinical Outcomes Assessments

Vineland Adaptive Behavior Scales were assessed as described in D’Souza et al.^5^, and identically to the phase 1/2 trial^6,7^. In short, there were six administrations of the Vineland 3^rd^ edition including at baseline and 180 (6 months), 365 days (1 year), 540 days (18 months), 730 days (2 years), and 1095 days (3 years) following gene transfer^6,7^. Here we report the baseline Adaptive Behavior Composite and the GSVs for the receptive communication, expressive communication, gross motor, and fine motor domains in accordance with the phase 1/2 trial^6^. Clinical global impression (CGI) evaluations were performed prospectively with a baseline CGI-severity (CGI-S), followed by five evaluations of CGI-improvement (CGI-I) or the change from baseline. CGI-I were scored during in-person follow-up evaluations which occurred at one month, 6-month, and 3 years following gene transfer. CGI-I were also scored via telemedicine at the 1 year and 2 year follow-up evaluations. CGI-S were scored between 1 (normal) and 7 (among the most extremely ill) and CGI-I were scored from 1 (very much improved) to 7 (very much worse) with 4 corresponding to no change. CGI scores were determined by a consensus among three researchers (PD, MTA, and CJT) based on their experience with GM1 and evaluations performed at the NIH^32^. All CGI were scored within two weeks of the clinical evaluation^6^.

### Neuroimaging

T1-weighted weighted imaging was acquired at 6 time points for this participant: twice prior to enrollment in the natural history study (ages 0-5 and 0-5), twice during the natural history study (ages 6-10 and 6-10), and three times during the expanded access trial (ages 6-10 [baseline] and 11-15). T1-weighted imaging acquisition for the natural history and gene therapy are identical and have been previously described^1,6,12,26,32^, all MRI imaging acquisition was performed on a Philips 3T system (Achieva, Philips Healthcare, Best, The Netherlands) under sedation or anesthesia as appropriate. A 3D T1-weighted protocol with a slice thickness of 1 mm, repetition time (TR) = 11 ms, echo time (TE) = 7 ms was used. DICOM images were converted to NIfTI using *dcm2niix*^43^. GT01 MRI scan acquisition prior to the natural history study (ages 0-5 and 0-5) are described in the Supplemental Appendix.

T1-weighted imaging was analyzed using volBrain’s *vol2Brain* to calculate volumes for the whole brain, lateral ventricles, and thalamus as previously described^1,6,44^. T1-weighted imaging was also analyzed using volBrain’s *BrainStructuresAges* to predict brain ages for the whole brain, lateral ventricles, and thalamus as previously described^12,29^. Brain structures age gap estimation (BSAGE) was calculated as the difference between the predicted brain age and participant age at each scan for each structure. Bilateral (thalamus and lateral ventricles) predicted ages and BSAGE measures were averaged. For this study, we report analysis of T1-weighted MRI scans up to three years following gene transfer in accordance with the efficacy endpoint of our previous investigation^6^.

Differential tractography was performed as previously described^6,13^; diffusion tensor imaging (DTI) was acquired during the same sedated session as the T1-weighted acquisition. DTI were acquired with the following parameters: TR = 6400 ms, TE = 100 ms, 15-gradient direction (30 directions for the natural history protocol), b-values 0 and 1000 s/mm^2^, slice thickness 2.5 mm, and FOV = 24 cm. GT01 had DTI acquisition at ages 6-10 and 6-10 (natural history) and ages 6-10, 11-15, and 11-15 during the expanded access study. DTI DICOM images were also converted to NIfTI using *dcm2niix*^13,43^. DTI were then preprocessed using MRtrix3’s (v3.0.4) *dwifslpreproc* command using MRtrix3’s *dwi2mask* followed by FSL’s (FSL, v6.0.5) *eddy* function^45–50^. Preprocessed DTI images were then analyzed in DSI Studio (DSI Studio, v November 6, 2023) where DTI images were quality checked for bad slices; a U-Net mask was created to remove non brain regions and generalized q-sampling imaging (GQI) reconstruction was performed with a diffusion sampling length ration of 1.25^6,13,51^.

We used differential tractography to compare changes in fractional anisotropy (FA) over time^6,13,35,52^. As described in Lewis et al.^6,13^, a baseline whole brain tractography was performed with an angular threshold of 60 degrees, a step size of one millimeter, one million seeds, a maximum length threshold of 200 millimeters, and a minimum length threshold of 30 millimeters. The fractional anisotropy (FA) map for each patient’s baseline scan was exported to an NIfTI file and utilized for differential fiber tractography. Whole brain differential tractography was calculated utilizing the same parameters as the baseline whole brain tractography using a 30 percent FA threshold. FA is a measure of diffusion anisotropy where increased FA indicates increased anisotropy and reflects increased myelination^35^. Follow-up scans were compared to the original baseline scan where fiber tract gains were determined in fiber tracts where the difference in FA between the follow-up scan and the baseline scan exceeded the FA threshold. Fiber tract losses were determined where the difference between the baseline scan and the follow-up scan exceeded the same threshold. Net fiber tract metrics for the number of fiber tracts and the volume of those tracts were determined as the difference between the fiber tract gains and fiber tract losses relative to the baseline whole brain tractography as a percentage change. Due to the sequence difference between the natural history DTI acquisition (30 gradient directions) and expanded access acquisition (15 directions) the two sets of scans were analyzed separately and compared as percentage changes relative to their respective baseline scans (age 6-10 and age 6-10).

## Acknowledgements

We thank GT01 and her family for the generosity of their time and efforts. We are also grateful to many staff members and care providers who contributed their expertise over the years. MRI analysis in this work utilized the computational resources of the Biowulf Linux cluster at the National Institutes of Health (http://hpc.nih.gov).

## Ethics Declaration

The Institutional Review Board at the NIH approved all three protocols for the natural history study, gene therapy clinical trial, and expanded access study (02-HG-0107, 19-HG-0101, and 19-HG-9958). Informed consent was completed with the parents and legal guardians of GT01. GT01 was assessed in her ability to provide assent and was not deemed capable.

## Data Availability

The data from this study is available from the corresponding author (CJT) at reasonable request.

## Authors’ Contributions

PD, JMJ, BJB, MS-E, HG-E, DRM, and CJT designed the study. CJL, HA, PD, JMJ, MTA, SA, AT, ZQ, MY, RYW, and CJT gathered the data; CJL, HA, GV, MB, SIC, AT, SA, XJ, ALG, AMK, TRF, HG-E, DRM, and CJT analyzed the data; CJL, MTA, XJ, ALG, AMK, and CJT vouch for the data and the analysis, CJL, PD, WAG, and CJT wrote the paper; CJL and CJT decided to publish the paper. The first draft of the manuscript was written by CJL.

## References

1. Lewis, C. J. et al. A Case for Automated Segmentation of MRI Data in Neurodegenerative Diseases: Type II GM1 Gangliosidosis. NeuroSci 6, (2025).

2. Sandhoff, R., Schulze, H. & Sandhoff, K. Ganglioside Metabolism in Health and Disease. Prog Mol Biol Transl Sci 156, 1–62 (2018).

3. Lang, F. M., Korner, P., Harnett, M., Karunakara, A. & Tifft, C. J. The natural history of Type 1 infantile GM1 gangliosidosis: A literature-based meta-analysis. Mol Genet Metab 129, 228–235 (2020).

4. Nicoli, E.-R. et al. GM1 Gangliosidosis-A Mini-Review. Front Genet 12, 734878 (2021).

5. D’Souza, P. et al. GM1 gangliosidosis type II: Results of a 10-year prospective study. Genet Med 26, 101144 (2024).

6. Lewis, C. J. et al. AAV9 Gene Therapy in Type II GM1 Gangliosidosis - A Phase 1-2 Trial. N Engl J Med 394, 1184–1194 (2026).

7. Sparrow, S. S., Cicchetti, D. V. & Saulnier, C. A. Vineland Adaptive Behavior Scales, Third Edition (Vineland-3). (Pearson, 2016).

8. Izumi, T., Ogawa, T., Koizumi, H. & Fukuyama, Y. Normal developmental profiles of CSF gangliotetraose-series gangliosides from neonatal period to adolescence. Pediatr Neurol 9, 297–300 (1993).

9. Kell, P. et al. A pentasaccharide for monitoring pharmacodynamic response to gene therapy in GM1 gangliosidosis. EBioMedicine 92, 104627 (2023).

10. Kaat, A. J. et al. Vineland-3 Growth Scale Values: Psychometric Properties for Clinical Trial Readiness in SCN2A. J Child Adolesc Psychopharmacol 35, 416–423 (2025).

11. Bonnefoy-Mazure, A., Coulon, G. D., Lascombes, P. & Armand, S. Follow-up of walking quality after end of growth in 28 children with bilateral cerebral palsy. Journal of Children’s Orthopaedics 14, 41 (2020).

12. Lewis, C. J. et al. Brain aging in Type II GM1 gangliosidosis. Commun Med (Lond) 10.1038/s43856-026-01745-w (2026) doi:10.1038/s43856-026-01745-w.

13. Lewis, C. J. et al. Differential tractography: an imaging marker for tissue degeneration in neurodegenerative diseases. Brain Commun 7, fcaf198 (2025).

14. Gray-Edwards, H. L. et al. 7T MRI Predicts Amelioration of Neurodegeneration in the Brain after AAV Gene Therapy. Mol Ther Methods Clin Dev 17, 258–270 (2020).

15. Gross, A. L. et al. Intravenous delivery of adeno-associated viral gene therapy in feline GM1 gangliosidosis. Brain 145, 655–669 (2022).

16. Weismann, C. M. et al. Systemic AAV9 gene transfer in adult GM1 gangliosidosis mice reduces lysosomal storage in CNS and extends lifespan. Hum Mol Genet 24, 4353–4364 (2015).

17. Gray-Edwards, H. L. et al. Novel Biomarkers of Human GM1 Gangliosidosis Reflect the Clinical Efficacy of Gene Therapy in a Feline Model. Molecular Therapy 25, 892 (2017).

18. McCurdy, V. J. et al. Sustained normalization of neurological disease after intracranial gene therapy in a feline model. Sci Transl Med 6, 231ra48 (2014).

19. Maurizi, N. et al. Incidence, timing, and clinical significance of adverse immune events after gene replacement therapy: A systematic review and meta-analysis. Mol Ther 34, 1340–1351 (2026).

20. Day, J. W. et al. Onasemnogene abeparvovec gene therapy for symptomatic infantile-onset spinal muscular atrophy in patients with two copies of SMN2 (STR1VE): an open-label, single-arm, multicentre, phase 3 trial. Lancet Neurol 20, 284–293 (2021).

21. Lek, A. et al. Death after High-Dose rAAV9 Gene Therapy in a Patient with Duchenne’s Muscular Dystrophy. N Engl J Med 389, 1203–1210 (2023).

22. Shieh, P. B. et al. Safety and efficacy of gene replacement therapy for X-linked myotubular myopathy (ASPIRO): a multinational, open-label, dose-escalation trial. Lancet Neurol 22, 1125–1139 (2023).

23. Shieh, P. B. et al. Learnings from Patient Mortality after Delandistrogene Moxeparvovec Administration: A Report of Two Cases and Expert Committee Considerations for Future Mitigation and Management. Hum Gene Ther 37, 262–279 (2026).

24. Donald, A. et al. Hemophagocytic Lymphohistiocytosis (Hlh)/Hyperinflammatory syndrome following high dose Aav9 therapy. American Society of Gene+ Cell Therapy (2025).

25. Butterfield, R. J. et al. AAV mini-dystrophin gene therapy for Duchenne muscular dystrophy: a phase 1b trial. Nat Med 31, 2712–2721 (2025).

26. Kolstad, J. et al. Natural history progression of MRI brain volumetrics in type II late-infantile and juvenile GM1 gangliosidosis patients. Mol Genet Metab 144, 109025 (2025).

27. Regier, D. S. et al. MRI/MRS as a surrogate marker for clinical progression in GM1 gangliosidosis. Am J Med Genet A 170, 634–644 (2016).

28. Nestrasil, I. et al. Distinct progression patterns of brain disease in infantile and juvenile gangliosidoses: Volumetric quantitative MRI study. Mol Genet Metab 123, 97–104 (2018).

29. Nguyen, H.-D., Clément, M., Mansencal, B. & Coupé, P. Brain structure ages-A new biomarker for multi-disease classification. Hum Brain Mapp 45, e26558 (2024).

30. Mhanna, H. Y. A., Oestreich, L. K. L., Vashistha, R., Wang, Y. & Vegh, V. Aging beyond diagnosis: the MRI brain age gap across disorders. Geroscience 10.1007/s11357-026-02315-7 (2026) doi:10.1007/s11357-026-02315-7.

31. Doering, E., Hoenig, M. C., Cole, J. H. & Drzezga, A. When Age Is More Than a Number: Acceleration of Brain Aging in Neurodegenerative Diseases. J Nucl Med 66, 1516–1521 (2025).

32. Lewis, C. J. et al. Retrospective assessment of clinical global impression of severity and change in GM1 gangliosidosis: a tool to score natural history data in rare disease cohorts. Orphanet J Rare Dis 20, 125 (2025).

33. Bingaman, A. et al. GM1-gangliosidosis: The caregivers’ assessments of symptom impact and most important symptoms to treat. Am J Med Genet A 191, 408–423 (2023).

34. Flotte, T. R. et al. AAV gene therapy for Tay-Sachs disease. Nat Med 28, 251–259 (2022).

35. Eichler, F. et al. Dual-vector rAAVrh8 gene therapy for GM2 gangliosidosis: a phase 1/2 trial. Nat Med 31, 2927–2935 (2025).

36. Laur, D. et al. Natural history of GM1 gangliosidosis-Retrospective cohort study of 61 French patients from 1998 to 2019. J Inherit Metab Dis 46, 972–981 (2023).

37. Strauss, K. A. et al. Onasemnogene abeparvovec for presymptomatic infants with three copies of SMN2 at risk for spinal muscular atrophy: the Phase III SPR1NT trial. Nat Med 28, 1390–1397 (2022).

38. Strauss, K. A. et al. Onasemnogene abeparvovec for presymptomatic infants with two copies of SMN2 at risk for spinal muscular atrophy type 1: the Phase III SPR1NT trial. Nat Med 28, 1381–1389 (2022).

39. Epstein, B. E. et al. Comparison of neonatal systemic and intracerebroventricular AAV9 gene therapy delivery demonstrating improved behavioral and phenotypic outcomes in a mouse model of Niemann-Pick disease, type C1. PLoS One 21, e0331275 (2026).

40. Leone, P. et al. Oligodendrocyte-targeted adeno-associated virus gene therapy for Canavan disease in children: a phase 1/2 trial. Nat Med 31, 3772–3779 (2025).

42. Leon-Astudillo, C. et al. Quantification and comparison of anti-AAV9 and anti-AAVrh74 antibodies in plasma and human milk: Implications for AAV-based gene therapy candidacy. J Neuromuscul Dis 12, 770–777 (2025).

43. Li, X., Morgan, P. S., Ashburner, J., Smith, J. & Rorden, C. The first step for neuroimaging data analysis: DICOM to NIfTI conversion. J Neurosci Methods 264, 47–56 (2016).

44. Manjón, J. V. et al. vol2Brain: A New Online Pipeline for Whole Brain MRI Analysis. Front Neuroinform 16, 862805 (2022).

45. Tournier, J.-D. et al. MRtrix3: A fast, flexible and open software framework for medical image processing and visualisation. Neuroimage 202, 116137 (2019).

46. Andersson, J. L. R. & Sotiropoulos, S. N. An integrated approach to correction for off-resonance effects and subject movement in diffusion MR imaging. Neuroimage 125, 1063–1078 (2016).

47. Smith, S. M. et al. Advances in functional and structural MR image analysis and implementation as FSL. Neuroimage 23 **Suppl 1**, S208–219 (2004).

48. Dhollander, T., Raffelt, D. & Connelly, A. Unsupervised 3-tissue response function estimation from single-shell or multi-shell diffusion MR data without a co-registered T1 image. in ISMRM workshop on breaking the barriers of diffusion MRI vol. 5 1 (Lisbon, 2016).

49. Jenkinson, M., Beckmann, C. F., Behrens, T. E. J., Woolrich, M. W. & Smith, S. M. FSL. Neuroimage 62, 782–790 (2012).

50. Woolrich, M. W. et al. Bayesian analysis of neuroimaging data in FSL. Neuroimage 45, S173–186 (2009).

51. Yeh, F.-C., Wedeen, V. J. & Tseng, W.-Y. I. Generalized q-sampling imaging. IEEE Trans Med Imaging 29, 1626–1635 (2010).

52. Yeh, F.-C. et al. Differential tractography as a track-based biomarker for neuronal injury. Neuroimage 202, 116131 (2019).

53. Kell, P. et al. Diagnostic and therapeutic applications of the glycan biomarker H3N2b in GM1 Gangliosidosis. Molecular genetics and metabolism 148, 110159 (2026).

