## Supplemental Appendix for "Expanded Access use of Intravenous Gene Transfer with AAV9-GLB1 in a Juvenile GM1 Gangliosidosis Participant"

**AAV9-GLB1 in a child with type II GM1 gangliosidosis**

**Supplementary Appendices**

Table of Contents

**Supplemental Methods**……………………………….……..………..……………………...……………*2*

GT01 Age 0-5 and 0-5 T1-weighted MRI Acquisition……………………………………………......……*2*

**Supplemental Results**………………………………….……..………..…………………………..………*3*

Adverse Events……………………………………………………………………………………………...*3*

Biochemical Testing………………………………………………………………………………………...*5*

Clinical Outcomes…………………………………………………………………………..………………*6*

Neuroimaging……………………………………………………………………………………………….*8*

**Supplemental Methods**

**GT01 Age 0-5 and 0-5 T1-weighted MRI Acquisition**

Age 0-5

T1-weighted structural images were acquired using a 3D T1 Gradient Echo (GR) sequence in the sagittal plane on a 3T Philips Achieva with an 8-channel SENSE head coil. Acquisition parameters were TR = 9.88 ms, TE = 4.59 ms, flip angle = 8°, slice thickness = 1 mm, acquisition matrix = 256 mm x 256 mm, and 164 sagittal slices.

Age 0-5

T1-weighted structural images were acquired using a 3D T1 MPRAGE sequence in the sagittal plane on a 3T Siemens Skyra scanner. Acquisition parameters were TR = 1800 ms, TE = 2.35 ms, flip angle = 8°, slice thickness = 0.9 mm, acquisition matrix = 256 mm x 256 mm, and 176 sagittal slices.

**Supplemental Results**

**Safety**

**Table S1. Cumulative AE’s During Study Period**

| **Event** | **Onset** | **CTCAE Grade^a^** | **AE Outcome^b^** | **Causality to Study Treatment^c^** |
| --- | --- | --- | --- | --- |
| 2 episodes of vomiting | Day -29 | Grade 2 Moderate | Resolved | Not related |
| Dehydration^*^ | Day -25 | Grade 3 Severe | Resolved | Not related |
| Infusion-related reaction (rituximab) | Day -21 | Grade 1 Mild | Resolved | Not related |
| Epistaxis | Day -13 | Grade 1 Mild | Resolved | Not related |
| Injection site reaction (Skin redness at PICC site) - Non-infectious ^*^ | Day -6 | Grade 1 Mild | Resolved | Not related |
| Hypotension | Day 0 | Grade 1 Mild | Resolved | Possibly related |
| Vomiting and decreased bowel sounds | Day 1 | Grade 2 Moderate | Resolved | Possibly related |
| Platelet count decreased | Day 7 | Grade 0^#^ | Resolved | Probably related |
| Oral mucositis | Day 36 | Grade 3 Severe | Resolved | Not related |
| Gastritis | Day 42 | Grade 2 Moderate | Resolved | Not related |
| Right upper eyelid stye (Eyelid function disorder) | Day 90 | Grade 1 Mild | Resolved | Not related |
| Dental caries | Day 107 | Grade 1 Mild | Resolved | Not related |
| Viral illness | Day 150 | Grade 1 Mild | Resolved | Not related |

^a^ Common Terminology Criteria for Adverse Events (CTCAE, v5.0)

^b^ Scored according to Common Terminology Criteria for Adverse Events (CTCAE) version 5.0.

^c^ As determined by the principal investigator.

^*^ Serious AE

^#^ Did not qualify as Grade 1 or higher on CTCAE

**Biochemical Testing**

Table S2. GT01 Biochemical Testing

| Day | AST | ALT | GGT | Platelets | D-Dimer | C3 | C4 |
| --- | --- | --- | --- | --- | --- | --- | --- |
| Day -30 | X | X | X | X |  |  |  |
| Day -21 | X | X |  | X |  |  |  |
| Day -14 | X | X |  | X |  |  |  |
| Day -7 | X | X |  | X |  |  |  |
| Day –1 | X | X | X | X |  |  |  |
| Day 0 |  |  |  |  |  |  |  |
| Day 1 |  |  |  |  |  |  |  |
| Day 2-3 | X | X |  | X |  |  |  |
| Day 4-6 |  |  |  | X | X | X | X |
| Day 7 | X | X | X | X | X |  |  |
| Day 14 | X | X | X | X |  |  |  |
| Day 21 | X | X | X | X |  |  |  |
| Day 30 | X | X | X | X |  |  |  |
| Day 45 | X | X | X | X |  |  |  |
| Day 60 | X | X |  | X |  |  |  |
| Day 75 | X | X | X |  |  |  |  |
| Day 90 | X | X | X | X |  |  |  |
| Day 180 | X | X | X | X |  |  |  |
| Day 210 | X | X | X |  |  |  |  |
| Day 270 |  |  |  |  |  |  |  |
| Year 1 | X | X | X | X | X | X | X |
| Month 18 | X | X | X | X |  |  |  |
| Year 2 | X | X | X | X |  |  |  |
| Year 3 | X | X | X | X |  | X | X |
| Year 4 |  |  |  |  |  |  |  |
| Year 5 | X | X | X | x |  |  |  |

**Clinical Outcomes**

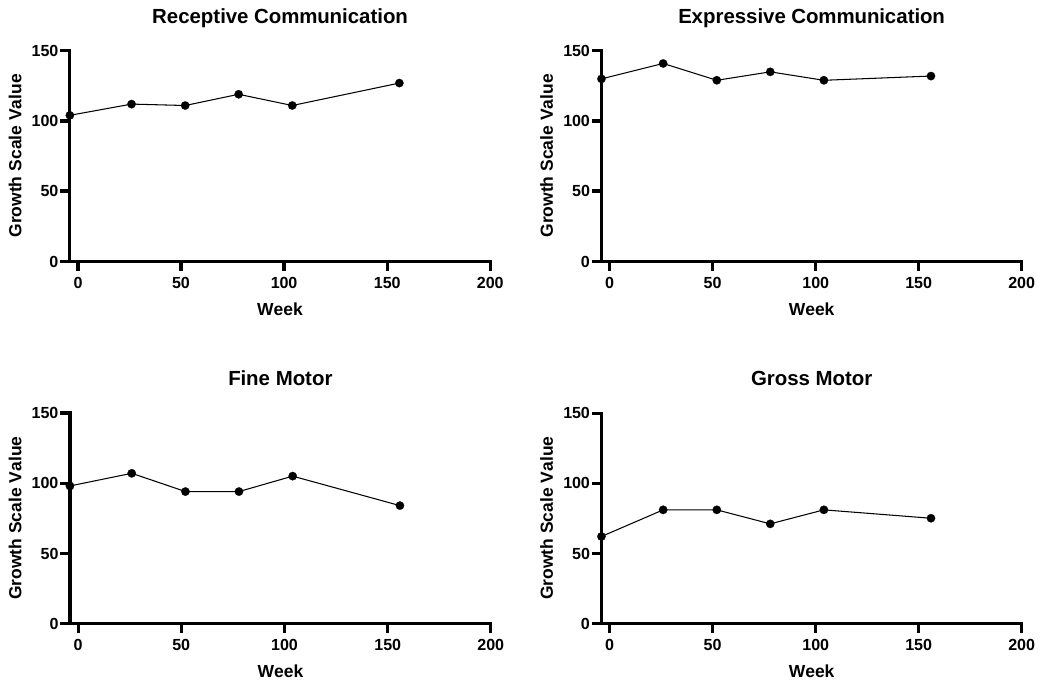

Figure S1. Vineland Growth Scale Values

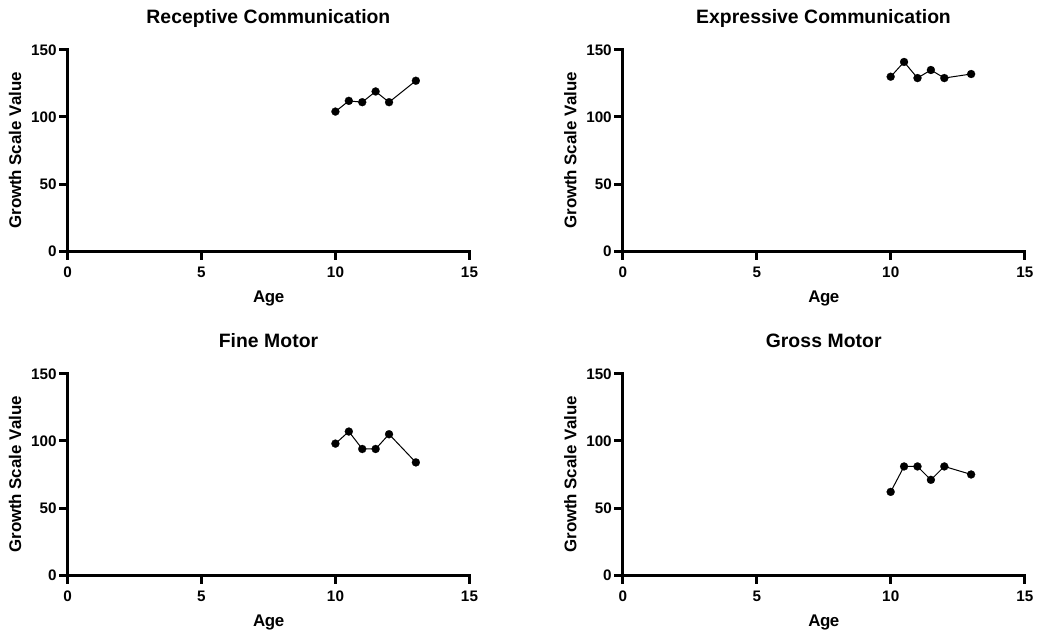

Figure S2. Vineland Growth Scale Values by participant age.

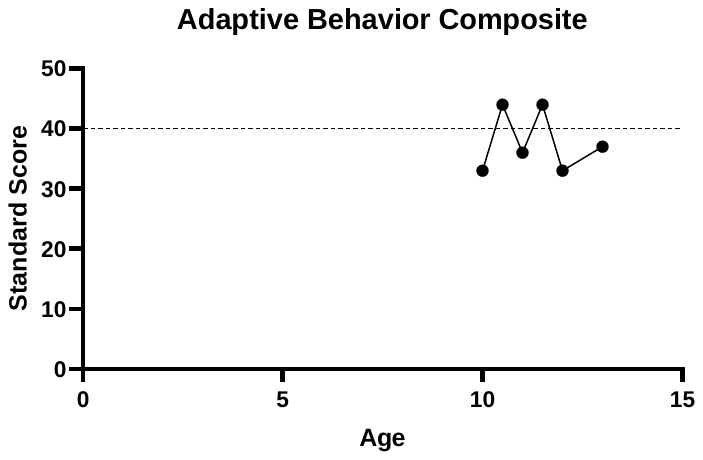

Figure S3. Vineland Adaptive Behavior Composite standard score. The dotted line is placed at a value of 40, which was the inclusion criteria for the phase 1/2 trial.

**Neuroimaging**

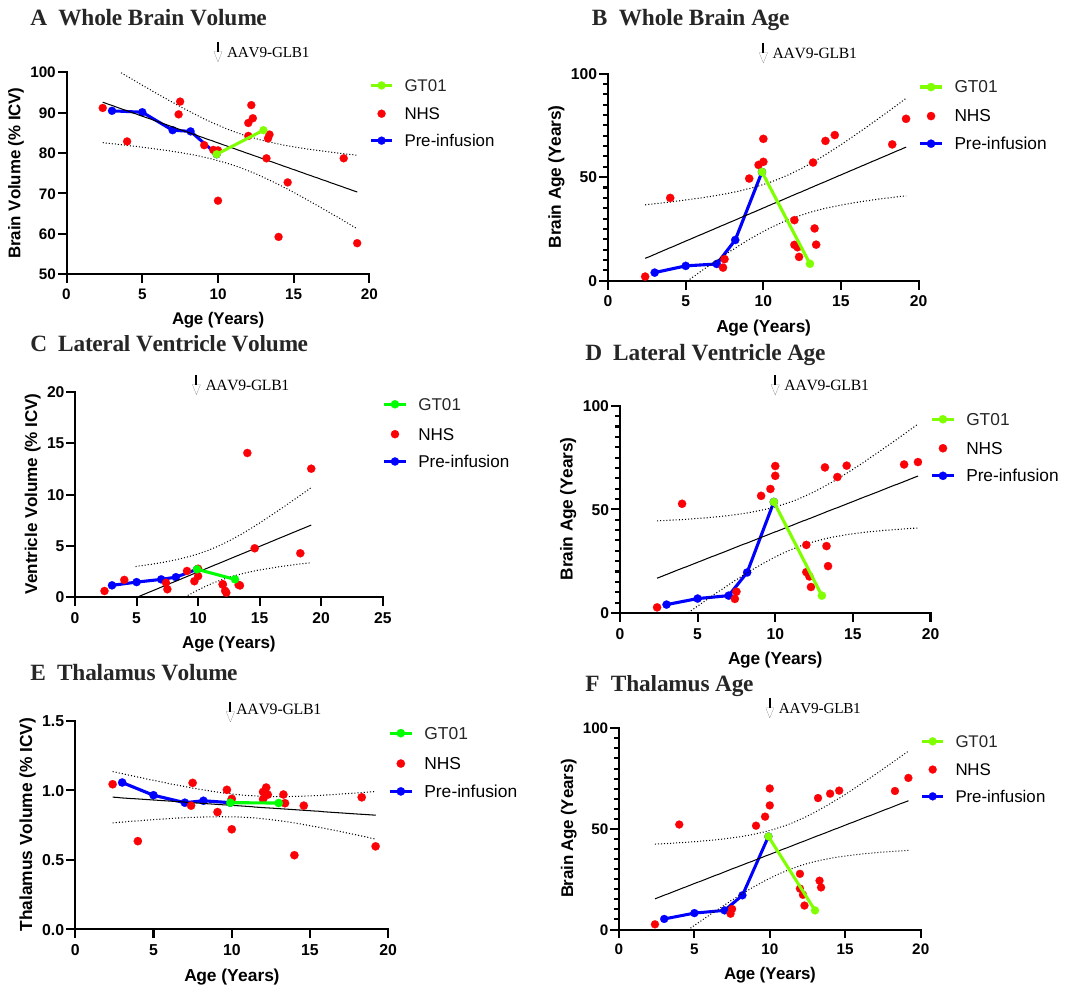

Figure S4. Brain MRI and Volumetric analysis. A.) GT01’s total brain volume as a percentage of total intracranial volume (ICV) from age 0-5 – 11-15 pre-infusion (blue) and post-infusion (green) in relation to the baseline scans from untreated natural history participants (red). B.) GT01’s predicted brain age for the whole brain from age 0-5 – 11-15 pre-infusion (blue) and post-infusion (green) in relation to the baseline scans from untreated natural history participants (red). C.) GT01’s lateral ventricle volume as a percentage of total intracranial volume (ICV) from age 0-5 – 11-15 pre-infusion (blue) and post-infusion (green) in relation to the baseline scans from untreated natural history participants (red). D.) GT01’s predicted brain age for the lateral ventricles from age 0-5 – 11-15 pre-infusion (blue) and post-infusion (green) in relation to the baseline scans from untreated natural history participants (red). E.) GT01’s thalamus volume as a percentage of total intracranial volume (ICV) from age 0-5 – 11-15 pre-infusion (blue) and post-infusion (green) in relation to the baseline scans from untreated natural history participants (red). F.) GT01’s predicted brain age for the thalamus from age 0-5 – 11-15 pre-infusion (blue) and post-infusion (green) in relation to the baseline scans from untreated natural history participants (red).
